# Cost-effectiveness analysis of Intravascular Temperature Management after cardiac arrest in England

**DOI:** 10.1101/2021.08.16.21262120

**Authors:** Mohsen Yaghoubi, Mehdi Javanbakht, Atefeh Mashayekhi, Mohsen Rezaei Hemami, Michael Branagan-Harris, Thomas R Keeble

## Abstract

**Background:** Although there is recognized that Targeted Temperature Management (TTM) can improve neurological outcomes and survival in patients resuscitated from out of hospital cardiac arrest, the cost-effectiveness of multiple methods of TTM is still uncertain.

**Objective:** This study aimed to evaluate the cost-effectiveness of Intravascular Temperature Management (IVTM) through Thermogard XP in targeted temperature management compared to surface cooling method after cardiac arrest in England.

**Methods:** We developed a multi-state Markov model that compared intravascular temperature management through Thermogard XP and the surface cooling method through two different devices including Blanketrol III and Arctic Sun 5000, over a short-term and lifetime horizon. Model input parameters were obtained from the literature and local databases. We assumed hypothetical cohort of 1,000 patients who need TTM after cardiac arrest per year in England. The outcomes were costs (in 2019 £) and quality-adjusted life-years (QALYs), discounted at 3.5% annually. Deterministic and probabilistic analyses were undertaken to examine the effect of alternative assumptions and uncertainty in model parameters on the results.

**Results:** In a simulated cohort of 1,000 patients who need TTM, the Thermogard XP resulted in direct cost savings of £2,339 and £2,925 (per patient) when compared with Blanketrol III and Arctic Sun 5000 respectively, and a gain of 0.98 QALYs over the patient lifetime. Total cost saving considering cohort of 1,000 patients is £2,339,479 and £ 2,925,109 when using IVTM through Thermogard XP compared with Blanketrol III and Arctic Sun 5000 respectively in life-time horizon. Results were robust against alternative assumptions, changes in values of input parameters, and alternative time horizons.

**Conclusion:** Implementation of intravascular temperature management using Thermogard XP can lead to cost-savings and improvement in patients’ quality of life versus surface cooling methods.

## Introduction

Cardiac arrest leads to loss of consciousness and death unless emergency resuscitation is given and the heart can be restarted^1^. Severe neurological injury has been considered as the main consequence of cardiac arrest following successful resuscitation^2^. Irreversible brain injury is the most common cause of death in the post-cardiac arrest phase^3^. Neurological damage happens not only during the cardiac arrest, but also during the reperfusion phase due to the generation of free radicals and other mediators^2^. Out of Hospital Cardiac Arrest (OHCA) affects 30,000 people each year in the United Kingdom (UK)^4^. 25% of OHCA population considered as an in-hospital cardiac arrest (IHCA) which hospitalized for therapy with a return of spontaneous circulation (ROSC)^4^ and around 84% of them who remain unconscious/comatose need targeted temperature management^5^.

Multiple methods of targeted temperature management are in clinical use to control and management of this patients ^6^. Different cooling methods have specific capabilities of extracting heat, which translate to different rates of achieving the intended target temperature. Methods of TTM may also differ in their ability to maintain a consistent target temperature as well as to control the rewarming phase after the Induced hypothermia (IH) protocol^7^. Mild hypothermia induced by surface cooling systems (SCS), intravascular temperature management (IVTM), and a combination of cooling methods have become standard therapy following cardiac arrest and it is recommended by the National Institute for Health and Care Excellence (NICE)^1^.

SCS work by circulating cold fluid or cold air through blankets or pads that are wrapped around the patient^7^. However, the IVTM provide precise temperature control during maintenance and rewarming phases of temperature management. ^2^ Thermogard XP^®^ has been used in the IVTM method to control a patient’s body temperature through central venous heat exchange^6^. It can be used to induce and maintain therapeutic hypothermia in critically ill patients after cardiac arrest^8^. The IVTM through Thermogard XP uses percutaneously placed central venous catheters, which can be placed in the subclavian, internal jugular, or femoral veins. Temperature control is achieved by circulating cool or warm saline in a closed loop through the catheter’s balloon^8^. The objective of this study was to estimate the cost and effectiveness of IVTM through Thermogard XP versus surface cooling methods as standard care in the hypothetical cohort of 1,000 patients who need TTM after cardiac arrest per year in England.

## Methods

### Model Overview

A de novo economic model was developed based on the current pathway for therapeutic hypothermia following cardiac arrest. The population was the hypothetical cohort of 1,000 patient (≥18 years) with ROSC after cardiac arrest and were admitted to critical care with ICD10 I460 and I469 as a primary or secondary diagnosis. The outcomes of interest included: improving neurological outcomes at hospital discharge and over a lifetime time horizon, long-term survival rates, reducing adverse events (AEs), total costs for each strategy, and incremental cost per quality-adjusted life year (QALY) gained. For each treatment arm, costs and outcomes were aggregated based on a series of decisions and events. The structure of the model was the same for the two treatment strategies. The recommended discount rate in the UK of 3.5% per annum for both costs and benefits^9^ was applied. As per NICE guidelines^10^, the base-case model considered all costs and health effects from the perspective of the UK NHS and personal social services (PSS). In order to fully capture the costs and benefits of Thermogard XP and the comparator(s), a lifetime time horizon was used in the base-case analysis.

### Intervention and comparator

The intervention in this study is the IVTM through Thermogard XP used in a hospital setting to control a patient’s body temperature through central venous heat exchange. This system circulates temperature-controlled saline within a closed-loop, multi-balloon intravascular catheter. The patient’s blood is cooled or warmed as it passes over the saline-filled balloons. No fluid is infused into or removed from the patient^8^. Recent European Society of Intensive Care Medicine Guidelines 2021: Post-resuscitation care ^11^, recommend TTM for adults after either OHCA or IHCA (with any initial rhythm) who remain unresponsive after ROSC. Body temperature is maintained at a constant value between 32 C and 36 C for at least 24 hours.

As comparators, in order to address heterogenicity in device costs, we considered Arctic Sun 5000 and Blanketrol III as two different devices based on the surface cooling methods (SCM). SCM has been considered as standard care based on the interventional procedure guidelines of therapeutic hypothermia following a cardiac arrest that has been developed by NICE^1^. According to this guideline, as soon as possible after the cardiac arrest, mild hypothermia is induced by using surface cooling methods such as heat exchange cooling pads, cooling blankets, ice packs. Core body temperature is maintained at 32–34°C for 12–24 hours from the start of cooling and is monitored using a bladder temperature probe or esophagus. Controlled re-warming is usually done over several hours^1^.

### Model structure

The economic model was developed in MS Excel for the hypothetical cohort of 1,000 patients who need therapeutic hypothermia after cardiac arrest per year in England.

A two-part economic model was developed, consisting of a short-term decision tree and a long-term Markov model. The decision tree has a relatively short horizon (2 months), and it estimates costs and effectiveness until the hospital discharge, and the Markov model used to follow individual patients with good and poor neurological outcomes over the analysis’ time horizon in annual cycles. The Markov model comprises of three health states that were used for survivors including good neurological outcome, poor neurological outcome, and death. The neurological outcome is defined based on the Cerebral Performance Category (CPC) as a validated scoring system for early stratification of neurological outcomes after cardiac arrest. In this category, CPC 1 and CPC 2 have been considered as a good neurological outcome, and a poor neurological outcome is defined as CPC 3 and CPC 4. Probabilistic neurological outcomes for the decision tree were determined at hospital discharge following the cardiac arrest event and were presumed to remain constant thereafter. Those who experienced different levels of neurological outcome enter the long-term survival Markov model, and they will remain in this health state until they die according to long-term mortality in terms of the level of neurological outcome. The model structure is illustrated in ***Figure1***.

**Figure 1:**
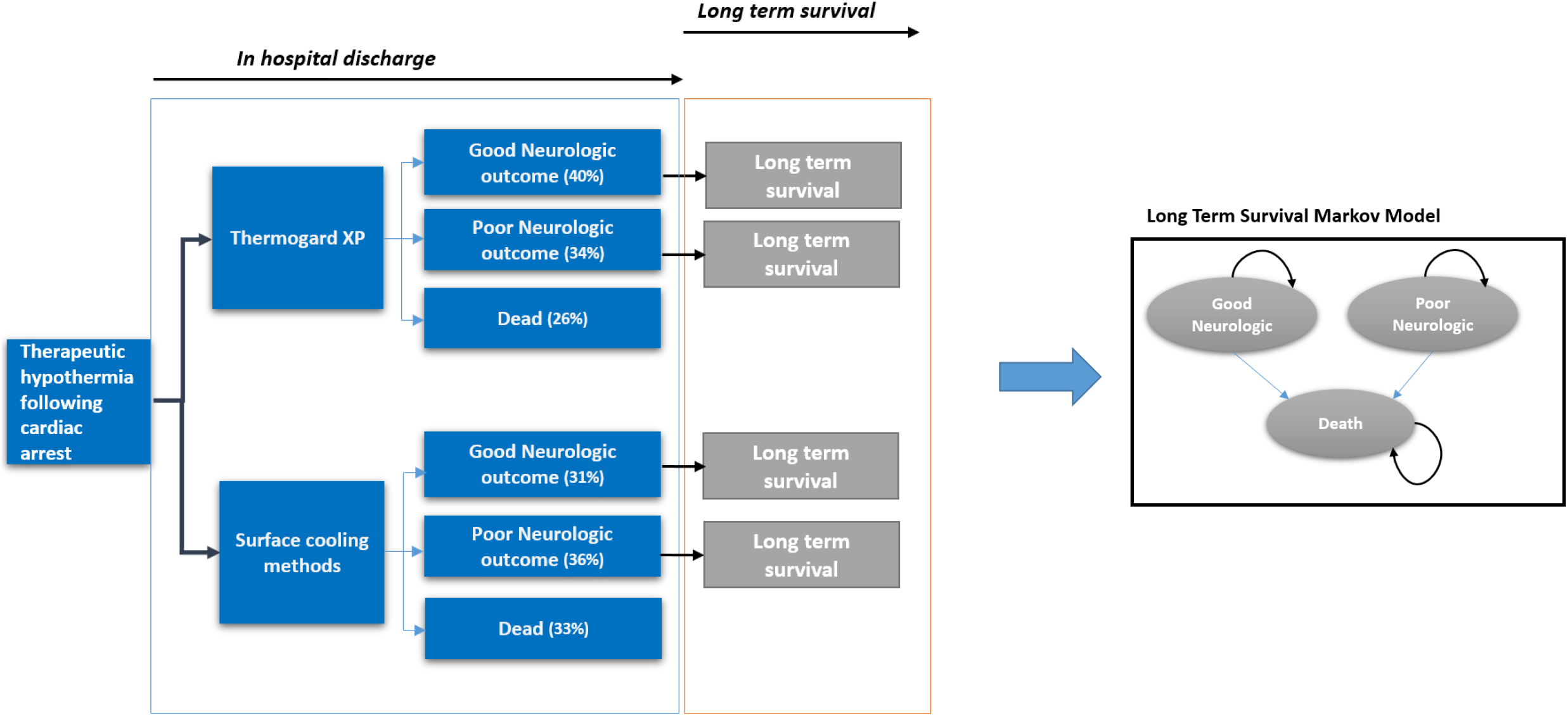
Economic model structure. **\***Percentage of good neurological outcome = (1-probability of poor neurological outcome derived from meta-analysis (see table 1); Percentage of poor neurological outcome = (1-mortality rate in each arm derived from meta-analysis) * probability of poor neurological outcome derived from meta-analysis (see table 1); Percentage of die= (mortality rate in each arm derived from meta-analysis * probability of poor neurological outcome derived from meta-analysis (see table 1)

### Model Inputs

#### Clinical efficacy

we conducted a subgroup meta-analysis on the studies which have been included in two recent review studies^6,12^ to estimate the pooled probability of unfavorable neurological outcomes and mortality over the hospital admittance duration. We estimated the pooled probability of a neurological outcome in intravascular temperature management versus surface cooling methods. Due to lack of evidence we assumed the same clinical efficacy for surface cooling method in terms of two different device in this group. To estimate the long-term mortality in terms of the level of neurological outcome, data from published Kaplan-Meier estimates for survival according to cerebral performance category score^13^ were digitalized and were used to estimate the parameters for parametric survival curves based on the methodology provided by Hoyle et al.^14^. Parametric survival models were fitted to the overall survival data to extrapolate the survival beyond the period of observation during the studies. Transition probabilities between the health states for the model were determined from these parametric survival functions. All estimated parametric survival functions are presented in *Appendix I*. All of the input parameters are presented in *Table 1*.

**Table 1:** Input parameters.

| Parameter | Value | Distribution | Source |
| --- | --- | --- | --- |
| ***Mortality rate (Hospital discharge)*** |  |  |  |
| Mortality rate (Thermogard XP) | 0.43 | Beta (4271,566.1) | Calculated using the pooled estimate reported in two recent meta-analysis<sup>6,12</sup> |
| Mortality rate (Surface cooling) | 0.48 | Beta (2643.2,2863.5) | Calculated using the pooled estimate reported in two recent meta-analysis<sup>6,12</sup> |
| ***Probability of poor neurological outcome*** |  |  |  |
| Probability of poor neurological outcome (Thermogard XP) | 0.60 | Beta (76.6,51.0) | Calculated using the pooled estimate reported in two recent meta-analysis<sup>6,12</sup> |
| Probability of poor neurological outcome (Surface cooling) | 0.69 | Beta (157.5,70.8) | Calculated using the pooled estimate reported in two recent meta-analysis<sup>6,12</sup> |
| ***Long-term survival*** |  |  |  |
| CPC1 | 25.70 | Weibull (0.08,0.76) | Phelps et al<sup>13</sup> |
| CPC2 | 8.11 | Weibull (0.10,0.86) | Phelps et al<sup>13</sup> |
| CPC3 | 2.74 | Weibull (0.19,0.72) | Phelps et al<sup>13</sup> |
| CPC4 | 0.61 | Weibull (0.53,0.59) | Phelps et al<sup>13</sup> |
| ***Health Utility*** |  |  |  |
| Good neurological outcome | 0.79 | Beta (8860.2,2355.2) | Stiell Ian et al, Hurdus et al and Fryback et al<sup>20–22</sup> |
| Poor neurological outcome | 0.39 | Beta (103.2,161.4) | Gage BF et al , Raina K et al<sup>23,24</sup> |
| In hospital | 0.74 | Beta (8260.1,2842.2) | Hurdus et al<sup>21</sup> |
| ***AE rates*** |  |  |  |
| Temperature Overshoot (Thermogard XP) | 0.15 | Beta (4.70,26.65) | Bartlett ES et al<sup>6</sup> |
| Shivering (Thermogard XP) | 0.25 | Beta (73.67,217.05) | Gillies et al<sup>25</sup> , Tomte et al<sup>26</sup> ,Deye et al<sup>27</sup> and Glover et al<sup>28</sup> |
| Serious bleeding (Thermogard XP) | 0.12 | Beta (1.90,13.95) | Bartlett ES et al<sup>6</sup> |
| Deep Venous Thrombosis (Thermogard XP) | 0.02 | Beta (15.06,737.89) | Bartlett ES et al <sup>6</sup> |
| Arrhythmia (Thermogard XP) | 0.15 | Beta (2.87,16.26) | Bartlett ES et al <sup>6</sup> |
| Pneumonia (Thermogard XP) | 0.56 | Beta (9.20,7.23) | Bartlett ES et al <sup>6</sup> |
| Temperature Overshoot (Surface cooling) | 0.33 | Beta (1.94,3.94) | Bartlett ES et al |
| Shivering (Surface cooling) | 0.28 | Beta (53.35,151.91) | Gillies et al<sup>25</sup> , Tomte et al<sup>26</sup> ,Deye et al<sup>27</sup> and Glover et al <sup>28</sup> |
| Serious bleeding (Surface cooling) | 0.07 | Beta (4.86,64.61) | Bartlett ES et al <sup>6</sup> |
| Deep Venous Thrombosis (Surface cooling) | 0.05 | Beta (22.81,433.38) | Bartlett ES et al <sup>6</sup> |
| Arrhythmia (Surface cooling) | 0.19 | Beta (6.16,26.28) | Bartlett ES et al <sup>6</sup> |
| Pneumonia (Surface cooling) | 0.48 | Beta (4.63,4.72) | Bartlett ES et al <sup>6</sup> |
| ***Costs (Device and consumable)*** |  |  |  |
| Device cost (Thermogard XP) | 34,648 | Fixed | NHS Supply Chain<sup>29</sup> |
| Annual cost of Thermogard XP | 4,717.87 | Fixed | Calculated* |
| Intravascular catheters (Cool Line) | 505.48 | Fixed | NHS Supply Chain<sup>29</sup><br>Icy= \$794.95<br>Quattro=\$822.0 |
| Start-up kit (model CG-500D) | 363.70 | Fixed | NHS supply chain |
| Foley temperature probe | 10.0 | Fixed | Medtech innovation briefing [MIB37]<sup>8</sup> |
| Temperature probe interface cable | 11.13 | Fixed | Medtech innovation briefing [MIB37]<sup>8</sup> |
| Device cost (Surface cooling-Blanketrol III) | 9,495.0 | Fixed | Medtech innovation briefing [MIB112]<sup>17</sup> |
| Annual cost of Blanketrol III | 1,469.0 | Fixed | Calculated* |
| Reusable connecting hose | 79.0 | Fixed | Medtech innovation briefing [MIB112]<sup>17</sup> |
| patient temperature probe cable | 40.0 | Fixed | Medtech innovation briefing [MIB112]<sup>17</sup> |
| Lite patient vest | 133.0 | Fixed | Medtech innovation briefing [MIB112]<sup>17</sup> |
| Lite adult | 129.0 | Fixed | Medtech innovation briefing [MIB112]<sup>17</sup> |
| Kool Kit | 347.0 | Fixed | Medtech innovation briefing [MIB112]<sup>17</sup> |
| Device cost (Surface Cooling-Arctic Sun 5000) | 20,600.0 | Fixed | Medtech innovation briefing [MIB112]<sup>17</sup> |
| Annual cost of Arctic Sun 5000 | 3,187.2 | Fixed | Calculated* |
| Temperature in cable | 116.3 | Fixed | Medtech innovation briefing [MIB112] |
| Temperature out cable | 93.09 | Fixed | Medtech innovation briefing [MIB112]<sup>17</sup> |
| Fill tube | 37.2 | Fixed | Medtech innovation briefing [MIB112]<sup>17</sup> |
| Fluid delivery line | 1,394.4 | Fixed | Medtech innovation briefing [MIB112]<sup>17</sup> |
| Drain tube | 32.5 | Fixed | Medtech innovation briefing [MIB112]<sup>17</sup> |
| Foley catheter temperature sensor | 349.1 | Fixed | Medtech innovation briefing [MIB112]<sup>17</sup> |
| Arctic gel pad kit | 628.3 | Fixed | Medtech innovation briefing [MIB112]<sup>17</sup> |
| Arctic gel pad | 545.9 | Fixed | Medtech innovation briefing [MIB112]<sup>17</sup> |
| Primary (Foley) | 10.3 | Fixed | Medtech innovation briefing [MIB112]<sup>17</sup> |
| Maintenance | 2,800.0 | Fixed | Medtech innovation briefing [MIB112]<sup>17</sup> |
| **Costs (Health care utilization)** |  |  |  |
| ICU costs per patient per day | 1,414.5 | Gamma (96,14.7) | NHS Reference Costs<sup>19</sup> |
| Hospitalization costs per patient per day | 354.0 | Gamma (96,3.7) | NHS Reference Costs<sup>19</sup> |
| Long term cost of CPC1 | 15,040.0 | Gamma (96,156.6) | Chan PS et al<sup>18</sup> |
| Long term cost of CPC2 | 23,993.0 | Gamma (96,249.8) | Chan PS et al<sup>18</sup> |
| Long term cost of CPC3 | 33,420.0 | Gamma (96,348) | Chan PS et al<sup>18</sup> |
| Long term cost of CPC4 | 24,587.0 | Gamma (96,256) | Chan PS et al<sup>18</sup> |
| **Cost (adverse event)** |  |  |  |
| Temperature Overshoot | 194.0 | Gamma (100,1.9) | NHS Reference Costs<sup>19</sup> |
| Shivering | 322.0 | Gamma (100,3.2) | NHS Reference Costs<sup>19</sup> |
| Serious bleeding | 752.0 | Gamma (100,7.5) | NHS Reference Costs<sup>19</sup> |
| Deep Venous Thrombosis | 862.0 | Gamma (100,8.6) | NHS Reference Costs<sup>19</sup> |
| Arrhythmia | 734.0 | Gamma (100,7.3) | NHS Reference Costs<sup>19</sup> |
| Pneumonia | 1,405.0 | Gamma (100,14.1) | NHS Reference Costs<sup>19</sup> |
| **Length of stay (LOS)** |  |  |  |
| Average LOS in ICU (days)-Good neurological | 8.2 | Gamma (12.9,0.6) | Petrie J et al<sup>16</sup> |
| Average LOS in a hospital ward (days)-Good neurological | 20.5 | Gamma (13.3,1.5) | Petrie J et al<sup>16</sup> |
| Average LOS in ICU (days)-Poor neurological | 21.0 | Gamma (67.8,0.3) | Petrie J et al<sup>16</sup> |
| Average LOS in a hospital ward (days)-Poor neurological | 65.0 | Gamma (19.3,3.4) | Petrie J et al<sup>16</sup> |
| Average LOS in ICU (days)-Non-survivor | 6.2 | Gamma (12.3,0.5) | Petrie J et al<sup>16</sup> |
| Average LOS in a hospital ward (days)-Non-survivor | 9.0 | Gamma (19.4,0.5) | Petrie J et al<sup>16</sup> |

*\*Annual cost of Thermogard XP calculated as: device cost divided by annualization factor (annualization factor estimated by considering lifetime (10 years) and discount rate (5%) for Thermogard XP and lifetime (8 years) for surface cooling device).*

#### Quality of life

To estimate the total QALYs gained in the Markov model, survival time was adjusted by health-related quality of life (HRQoL). The utility values/quality of life inputs include utilities of health states and the disutility associated with treatment-related adverse events. HRQoL of the cardiac arrest survival based on patient’s CPCs has been derived from relevant published literature and previous economic evaluation studies in this area^15^ and are presented in *Table 1*. According to the literature, we assumed a consistent utility following survival after cardiac arrest^16^.

#### Adverse events

We derived the risk difference in adverse events in intravascular temperature management groups versus surface cooling methods based on the result of a recent random-effect meta-analysis study^6^. The risk difference in occurrence of adverse events, along with the disutility of each event, are presented in *Table 1*.

#### Costs

The following costs were included: annual capital cost and consumable cost of Thermogard XP and surface cooling system, the cost associated with a stay in the intensive care unit (ICU), hospital ward and costs associated with the treatment of AEs. All cost input parameters are reported in *Table 1*. The cost of Thermogard XP was extracted from Medtech innovation briefings developed by NICE^8^ and NHS supply chain, and the cost of the surface cooling methods were provided in different Medtech innovation briefings developed by NICE^17^. Health care resource utilization costs in terms of neurological outcome, along with length of stay, were derived from a UK single-center cost study that estimated the hospital costs of out-of-hospital cardiac arrest patients treated in intensive care evaluation using the national tariff-based system^16^. The long-term cost of poor and good neurological outcomes was derived from a US-based study^18^. AE costs were obtained from NHS reference costs^19^ and these were adjusted by the rate of each AE in the two arms. Costs were measured in UK pound sterling (£) for a 2019 price year.

#### Analysis

The cumulative estimates of costs and effectiveness were reported for each treatment arm. The incremental cost per life-year and incremental cost per QALY was reported. Deterministic sensitivity analyses were conducted to investigate the impact of key assumptions and parameter values used in the base-case analysis. The results were reported using Tornado diagrams and supporting tables. The model allows for probabilistic sensitivity analysis (PSA) using Monte Carlo simulation. This form of analysis is used to estimate parameter uncertainty. To conduct the PSA, probabilistic distributions that have been assigned to each input in the model were used to randomly select new plausible values. Each new sampled value applied in the model and the new results of the model were recorded. This process was repeated for a large number of iterations (10,000) to produce a distribution of results from the model. The outcomes were reported using cost-effectiveness scatter plots and cost-effectiveness acceptability curves (CEACs).

## Results

### Meta-analysis

The result of the subgroup meta-analysis on neurological outcome (*Figure 2)* indicates that seven studies^25,30–35^ compared the neurological outcome in intravascular temperature management versus the surface cooling method in hospital discharge duration. Intravascular temperature management showed a lower probability of unfavorable neurological outcomes than surface methods (OR 0.73 [95% CIs 0.61–0.88]). As shown in *Figure 3*, twelve studies ^25,30–40^ compared the mortality in intravascular temperature management versus surface cooling method in hospital discharge duration. Intravascular temperature management showed a lower probability of mortality than surface methods (OR 0.87 [95% CIs 0.75–1.02]).

**Figure 2:**
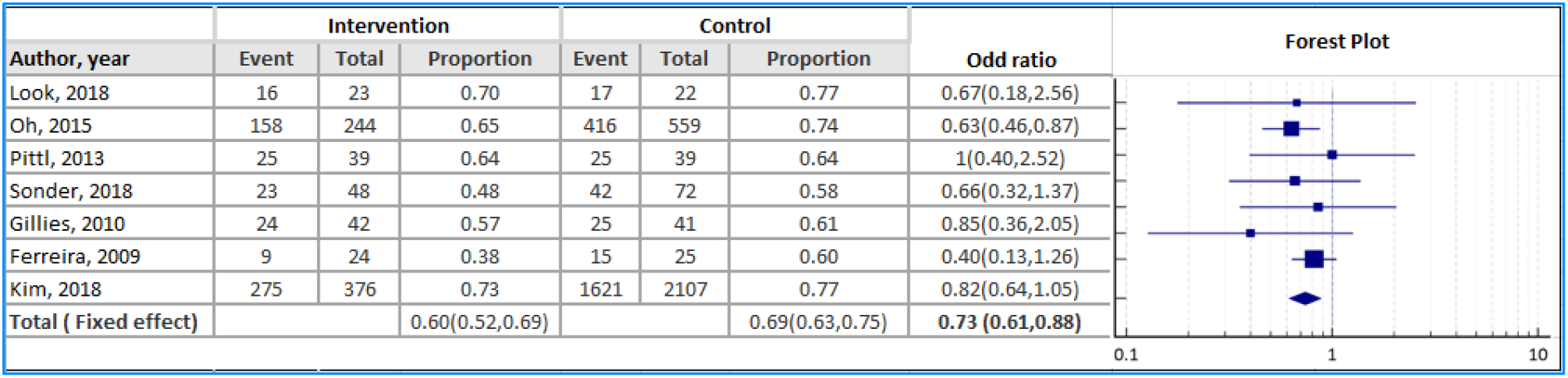
Forest plot of poor neurological outcome in hospital discharge duration: intravascular temperature management vs. surface methods

**Figure 3:**
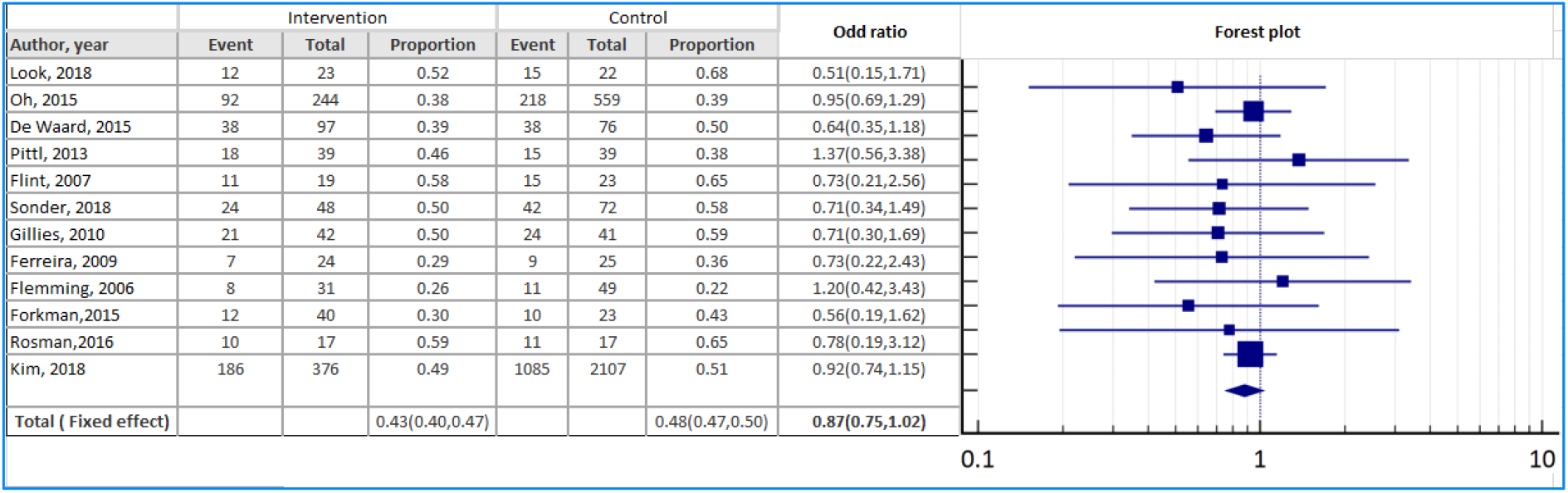
Forest plot of mortality in hospital discharge duration: intravascular temperature management vs. surface methods

### Cost-effectiveness analysis

The result of base-case cost analysis over a life-time horizon demonstrated that the average device cost per patient (annual cost of equipment along with consumable cost) of Thermogard XP was £932.91 per patients compared to £503.4 for Blanketrol III and £1075.3 for Arctic Sun 5000. While cost of ICU stay in treatment with IVTM through Thermogard XP was £17,108 per patients compared with £17,204 in surface cooling methods. Considering the cost of hospital ward and adverse event treatments, the total average discounted cost for intervention was £82,846. In comparison, the average discounted cost for Blanketrol III and Arctic Sun 5000 was £85,185 and £85,771 respectively. Treatment with Thermogard XP led to an increase of 0.98 in discounted QALYs relative to Blanketrol III and Arctic sun 500 over a lifetime time horizon *(Table 2)*. The estimated incremental cost-effectiveness ratio (ICER) was dominant for Thermogard XP versus Arctic Sun 5000 and Blanketrol III over a lifetime horizon, i.e., the intervention was less costly and more effective than the comparator(s).

**Table 2:**
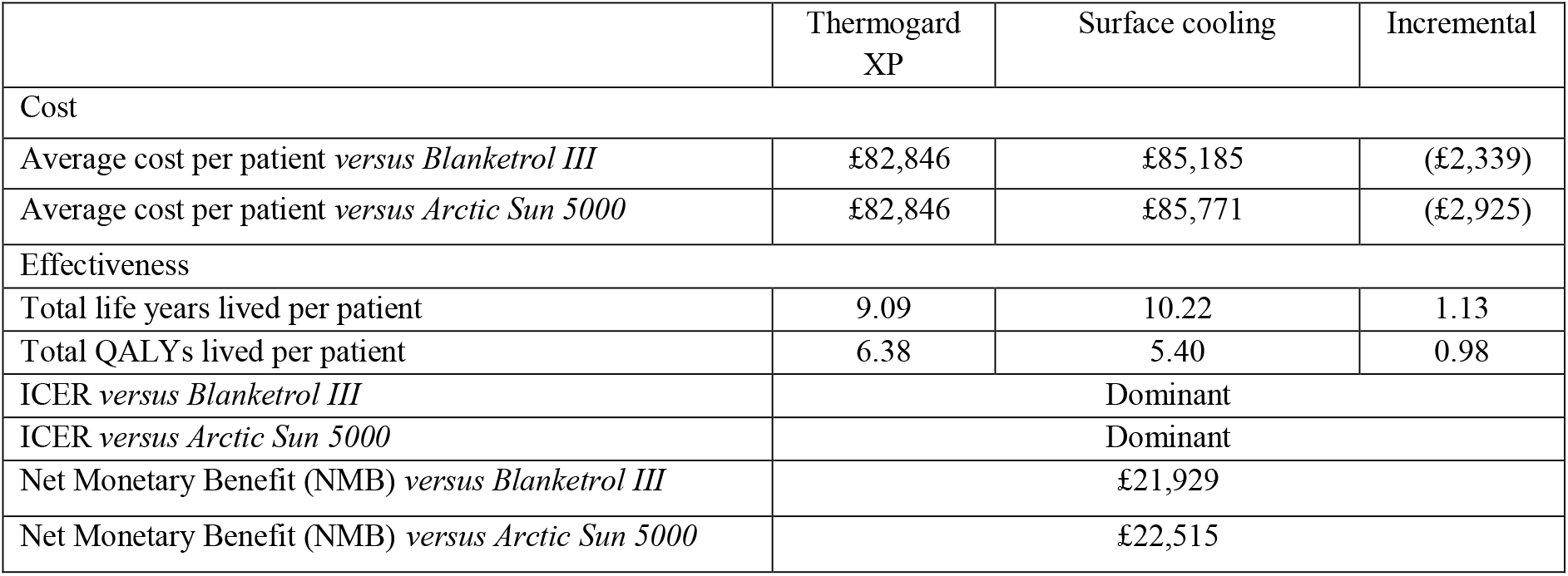
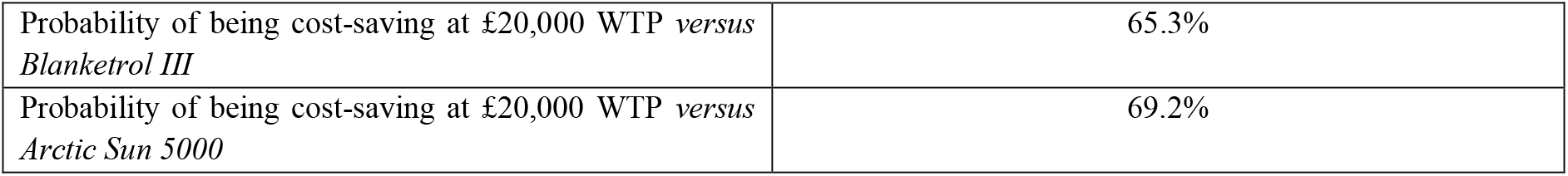
Result of cost-effectiveness analysis over lifetime time horizon

|  | Thermogard XP | Surface cooling | Incremental |
| --- | --- | --- | --- |
| Cost |  |  |  |
| Average cost per patient *versus Blanketrol III* | £82,846 | £85,185 | (£2,339) |
| Average cost per patient *versus Arctic Sun 5000* | £82,846 | £85,771 | (£2,925) |
| Effectiveness |  |  |  |
| Total life years lived per patient | 9.09 | 10.22 | 1.13 |
| Total QALYs lived per patient | 6.38 | 5.40 | 0.98 |
| ICER *versus Blanketrol III* | Dominant |  |  |
| ICER *versus Arctic Sun 5000* | Dominant |  |  |
| Net Monetary Benefit (NMB) *versus Blanketrol III* | £21,929 |  |  |
| Net Monetary Benefit (NMB) *versus Arctic Sun 5000* | £22,515 |  |  |

|  |  |
| --- | --- |
| Probability of being cost-saving at £20,000 WTP *versus Blanketrol III* | 65.3% |
| Probability of being cost-saving at £20,000 WTP *versus Arctic Sun 5000* | 69.2% |

Results of the probabilistic sensitivity analysis, as shown in *Figures 4-1 and 4-2*, are presented in the form of a cost-effectiveness plane and CEAC plot respectively for IVTM through Thermogard XP compared with surface cooling methods. Visualization of the results in the cost-effectiveness plane demonstrated that most of the times the incremental cost-effectiveness rate was located in the cost-effective and cost-saving quadrants. Assuming a willingness-to-pay threshold for a QALY of £20,000, the probability of being cost-saving for IVTM through Thermogard XP would be 69.2 % and 65.3% versus the Arctic Sun 5000 and Blanketrol III based on the CEAC plot *(Figure 4-1 and 4-2)*.

**Figure 4-1:**
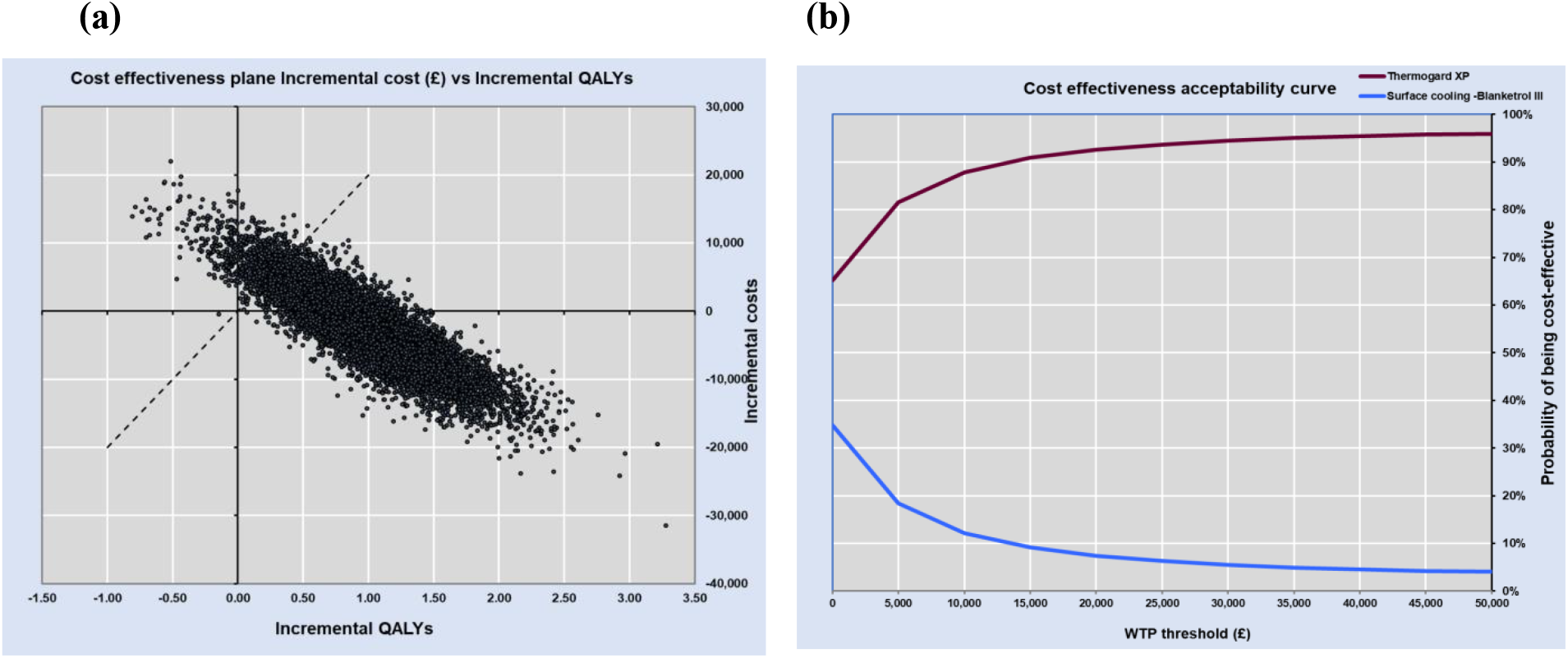
cost-effectiveness plane (a) and cost-effectiveness acceptability curve plot (b) for IVTM through Thermogard XP versus Blanketrol III

**Figure 4-2:**
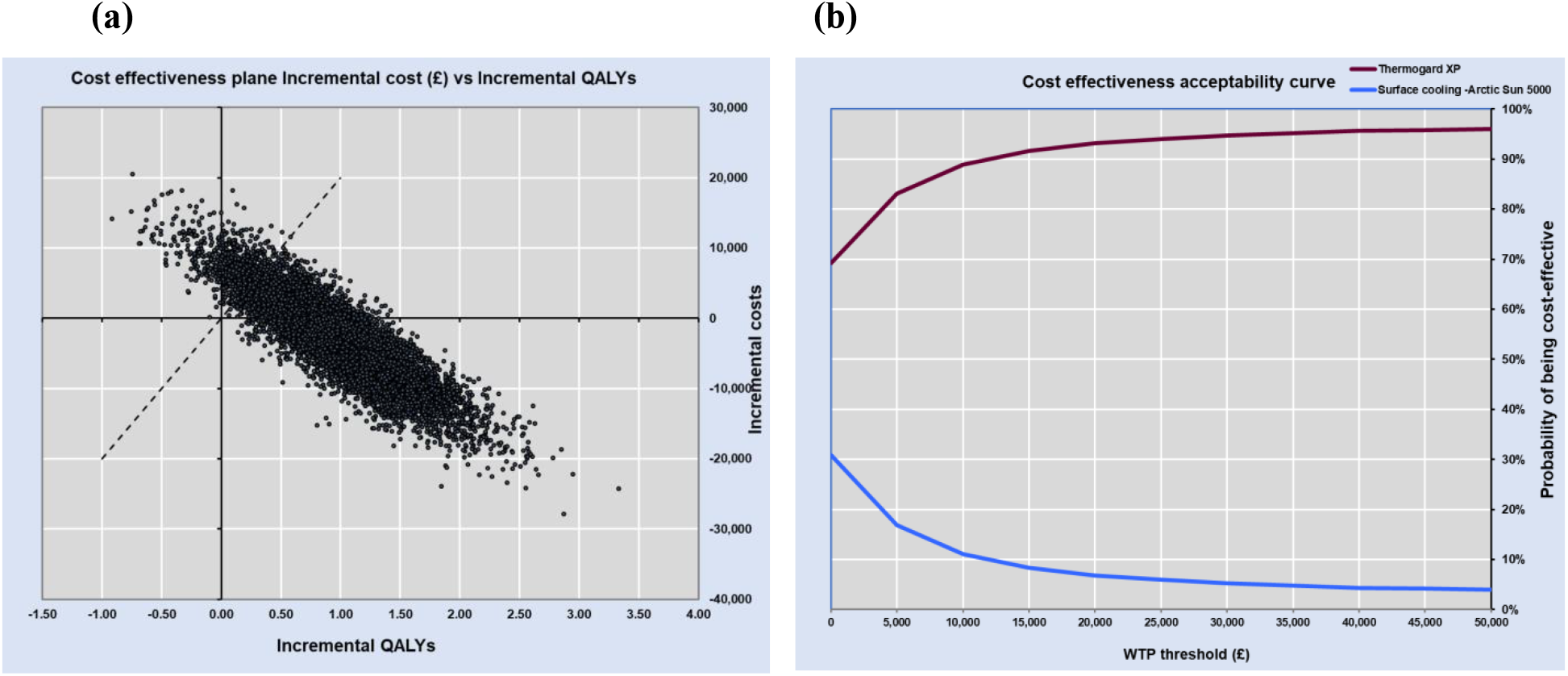
cost-effectiveness plane (a) and cost-effectiveness acceptability curve plot (b) for IVTM through Thermogard XP versus Arctic Sun 5000

In the deterministic sensitivity analysis, key cost and outcome parameters were subject to hypothetical increases or decreases of 25% to determine the key drivers of the model results. Results from the one-way sensitivity analyses generally supported the base-case findings, and findings from the different scenario analyses are shown in *Figures 5-1 and 5-2* in terms of change in incremental cost versus Blanketrol III and Arctic Sun 5000 respectively. Regarding the result of deterministic sensitivity analysis, the probability of neurological outcome had the most significant effect on the base-case results. The results of the base-case analysis over the hospital discharge horizon are presented in *Appendix II*.

**Figure 5).**
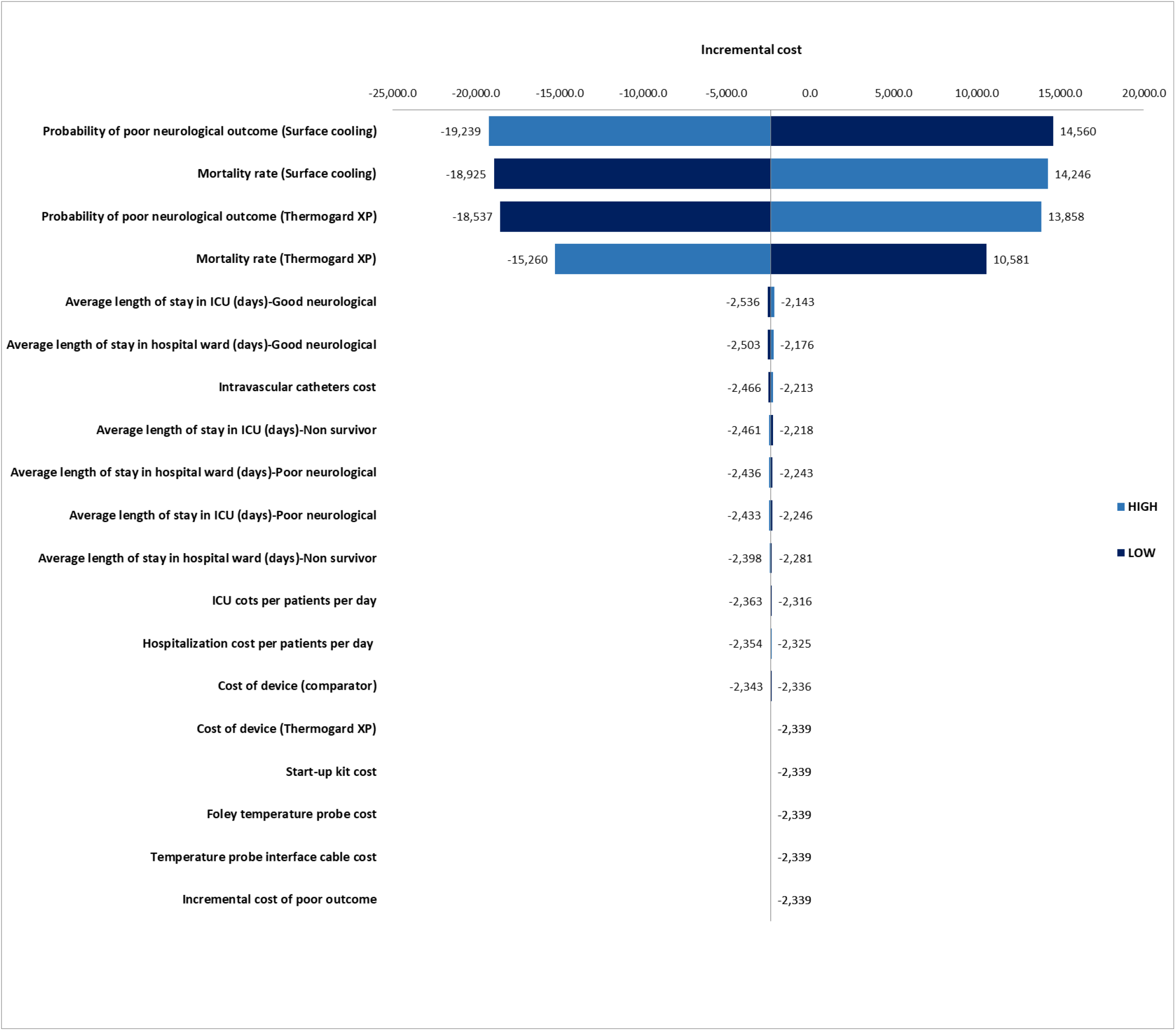
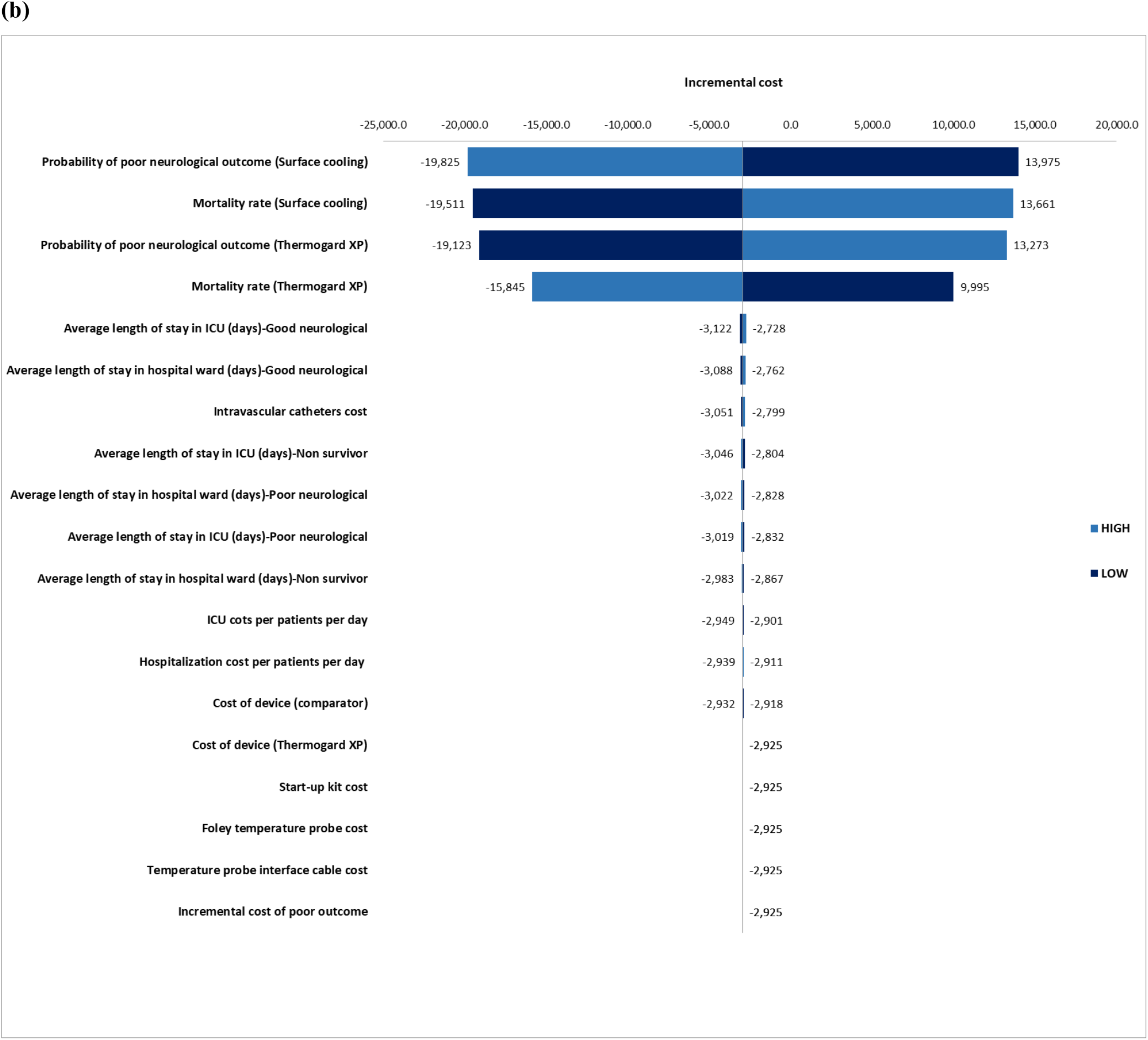
Tornado Diagram for change in Incremental cost versus Blanketrol III (a) and Arctic Sun 5000 (a)

## Discussion

We performed a cost-effectiveness analysis of intravascular temperature management by Thermogard XP compared with the surface cooling method by Blanketrol III and Arctic Sun 5000. Results of the cost-effectiveness analysis over a lifetime time horizon indicated that this intervention is dominant compared with the surface cooling method. To the best of our knowledge, this is the first cost-effectiveness analysis to evaluate IVTM using a Markov model.

According to the literature, therapeutic hypothermia following cardiac arrest leads to improved neurological and survival outcomes. Although different temperature management procedures are cost-effective to control cardiac arrest survival^15,41^, identifying the most cost-effective cooling procedure still needs to be explored. Since temperature is one of the four main vital signs, achieving the optimal temperature management procedure should be considered when treating many critically ill or surgical patients.

A previous study determined that therapeutic hypothermia with a cooling blanket among cardiac arrest survivors improves clinical outcomes and is a cost-effective intervention versus conventional care in the United States^15^. In this study, patients receiving therapeutic hypothermia gained an average of 0.66 quality-adjusted life years compared with conventional care, at an incremental cost of $31 254. Another study evaluated different methods of temperature management including blanket cooling, peritoneal lavage, and veno-venous extracorporeal membrane oxygenation V–V ECMO^41^. The result of this study showed that a cooling blanket is the most cost-effective intervention with an ICER of $58,329/QALY.

Since recent review studies^6^ determined that IVTM was associated with improved neurological outcomes compared to surface cooling methods among survivors resuscitated following cardiac arrest, conducting an economic evaluation to assess the costs and outcomes of IVTM method is important to help health care policymakers to identify the optimal procedure. Beside improving neurological outcome and survival rate, IVTM is associated with reduce the burden of some significant adverse event including temperature overshoot, Arrhythmia, deep venous thrombosis and shivering. These complications will complicate the treatment process and delay hospital discharge which will impose more costs to the health care system. For instance, shivering is a common side effect in targeted temperature management and can lead to cerebral and metabolic stress ^28,42^ that need to additional usage of sedation, more length of stay and increase the burden of health care utilization. Recent finding shows that IVTM is associated with reduce rate of shivering as 2.36% compared to surface cooling that should be consider as clinical and economic impact of IVTM.

Furthermore, some important clinical outcome of IVTM including the short time to resuscitation and rapid rewarming period led to lower temperature variability and slower rewarming rate during the maintenance stage.^43,44^ These effects might be significant particularly in patients who receive sedative and analgesics medication in surface cooling method and conclude that, IVTM in addition to saving health care resource utilization, it also has favorable clinical effect.

In this study, we found that the IVTM method is likely to be the most cost-effective strategy among current temperature management procedures for treating hypothermia after cardiac arrest. The probability of poor neurological outcome has been considered as the main driver in this analysis, based on deterministic sensitivity analysis. There is a significant difference between the length of stay in ICU and hospital ward in terms of poor and good neurological outcome in the UK health care system. As a result, the probability of neurological outcome parameters has a significant effect on costs, clinical outcomes, and results of the economic evaluation.

The strength of our study is that we used multiple sources of high-level evidence on the probability of a neurological outcome in IVTM and the surface cooling method. Also, the incorporation of uncertainty in model inputs and assumptions into the results should provide reassurance that the overall findings are not materially affected by uncertain evidence. The limitations of our study should also be acknowledged. Due to lack of evidence, we assumed that the status of neurological outcome among survivors of cardiac arrest is constant over a lifetime time horizon and also, we combined CPC 1 and CPC 2 as good and CPC 3 and CPC 4 as poor neurological outcome status, which did not allow for variation in long-term care costs between all different neurological states. To model this assumption, we derived the rate of each neurological status including CP1, CP2, CP3, and CP4 ^13^ and we then estimated the weighted average hazard ratio by adjusting the rate of CPC1 and CPC 2 for good and CPC3 and CPC 4 for the poor neurological outcome. Also, we just assigned the incremental cost of poor neurological outcome among patients in this condition until death to avoid over-estimation of poor outcome costs in the long-term. Moreover, the main source published Kaplan-Meier estimates for survival according to cerebral performance category score and rate of each status was a US-based study^13^ and we used a most recent UK national life table^45^ to estimate age and sex-adjusted hazard ratio for the final survival parameters in our model.

## Conclusion

Our findings show that using the intravascular temperature management is associated with better neurological outcome and survival rate in both short term and long-term horizon. In addition, intravascular temperature management improves life-year gains and reduces the total cost per patient over a lifetime time horizon versus the surface cooling method for managing patients’ temperature after cardiac arrest.

## Data Availability

The authors confirm that the data supporting the findings of this study are available within the article and its supplementary materials.

## Author contributions

MJ conceived the study question. MY and MJ, MRH developed the analysis plan, conducted the economic model, and drafted the manuscript. All authors contributed to the study design and provided input on the model, read, and approved the final draft of the manuscript.

## Availability of data and material

Not applicable

## Appendix

### Appendix I Parametric survival function estimation

All data from published Kaplan-Meier curves from Phelps and et al.^13^ were digitalized and were used to estimate the parameters for the parametric survival curves. Parametric survival models were fitted to the OS data to extrapolate the survival of patients beyond the period of observation during the studies. Specifically, the following survival distributions were considered: exponential, Weibull, logistic, log-logistic, and log-normal. The goodness-of-fit criteria, including the Akaike information criterion (AIC) and the Bayesian information criteria (BIC), were estimated for each parametric function.

*Figures A1-A4* below, present the estimated OS curves for each survival distribution in terms of four levels of Cerebral Performance Category (CPC). Because the Weibull distribution has lower AIC and BIC values, OS was extrapolated using this distribution. All estimated parametric Weibull survival functions are presented in *Table A1*.

Since we have combined CPC1 and CPC 2 as a good neurological outcome and CPC3 and CPC 4 as a poor neurological outcome, we used the rate of each neurological status including CP1, CP2, CP3, and CP4 from this study and then we incorporated the weighted mean of the hazard ratio for the good neurological outcome by adjusting the survival parameter and rate of CPC1 and CPC2 and we applied the same approach for the poor neurological outcome by adjusting the survival parameters and the rates of CPC3 and CPC 4 for the poor neurological outcome. Finally, we used the UK national life table to estimate the age and sex-adjusted hazard ratio for the good and poor neurological outcome.

**Figure A1:**
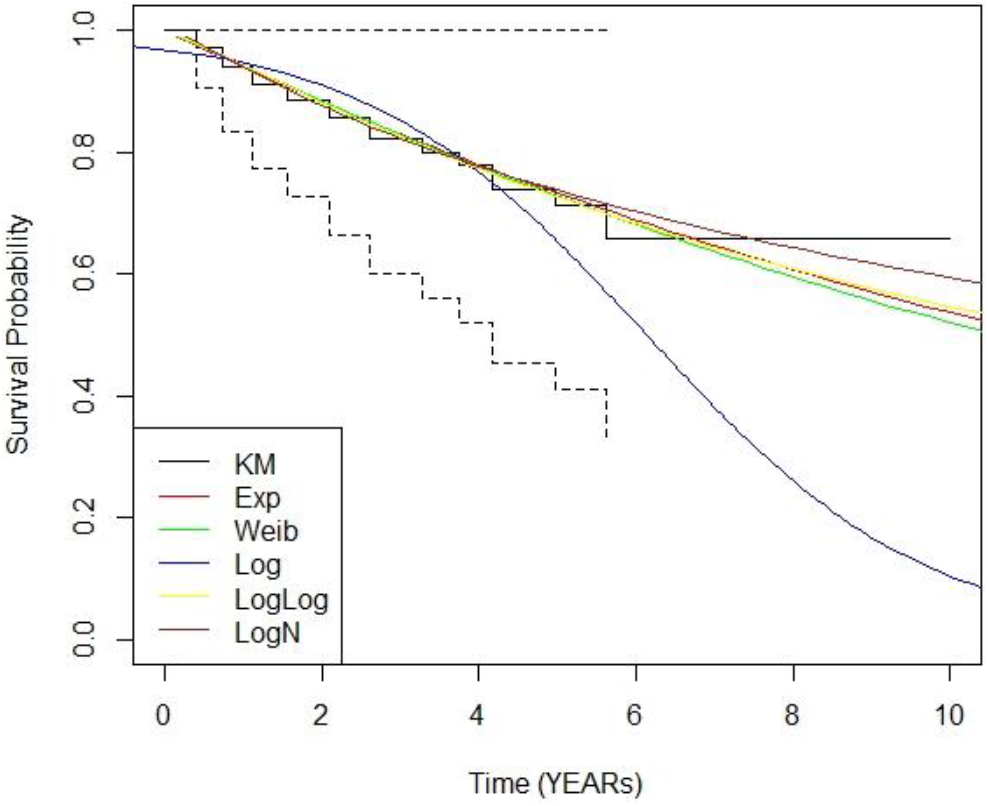
Estimated OS curves for the CPC 1 using different distributions.

**Figure A2:**
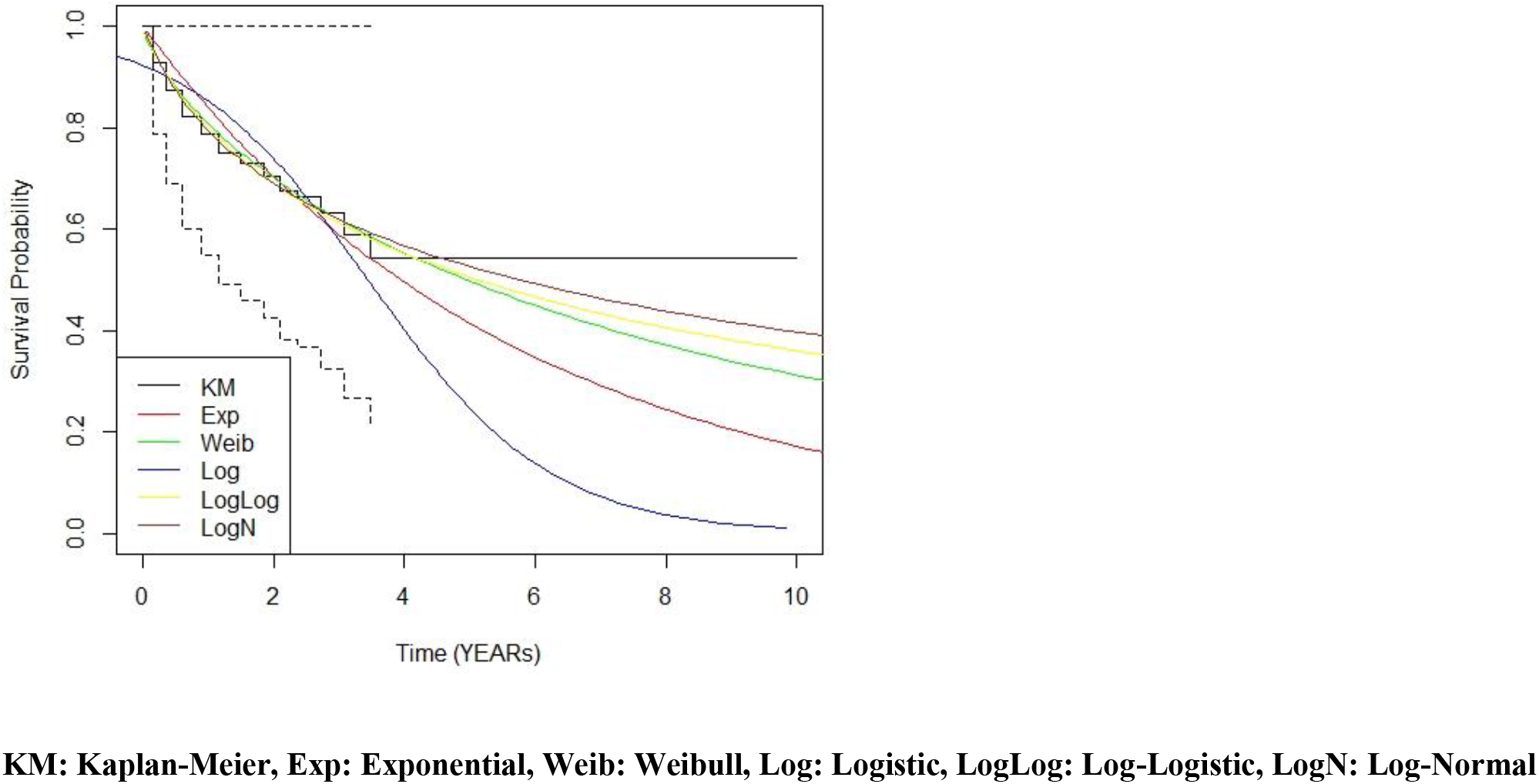
Estimated OS curves for the CPC 2 using different distributions.

**Figure A3:**
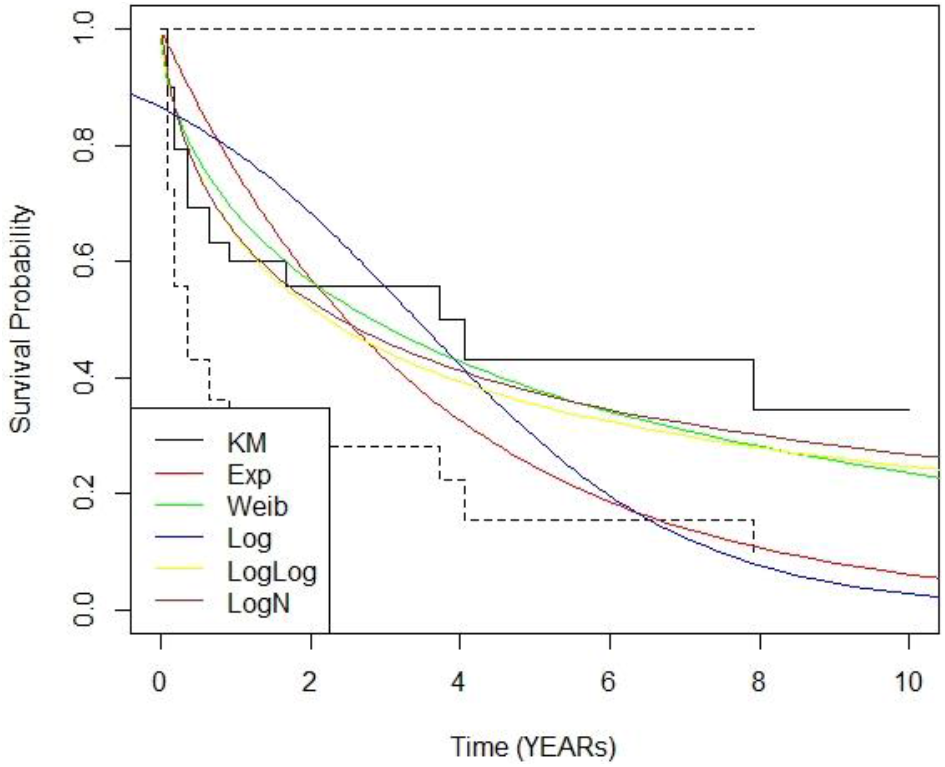
Estimated OS curves for the CPC 3 using different distributions.

**Figure A4:**
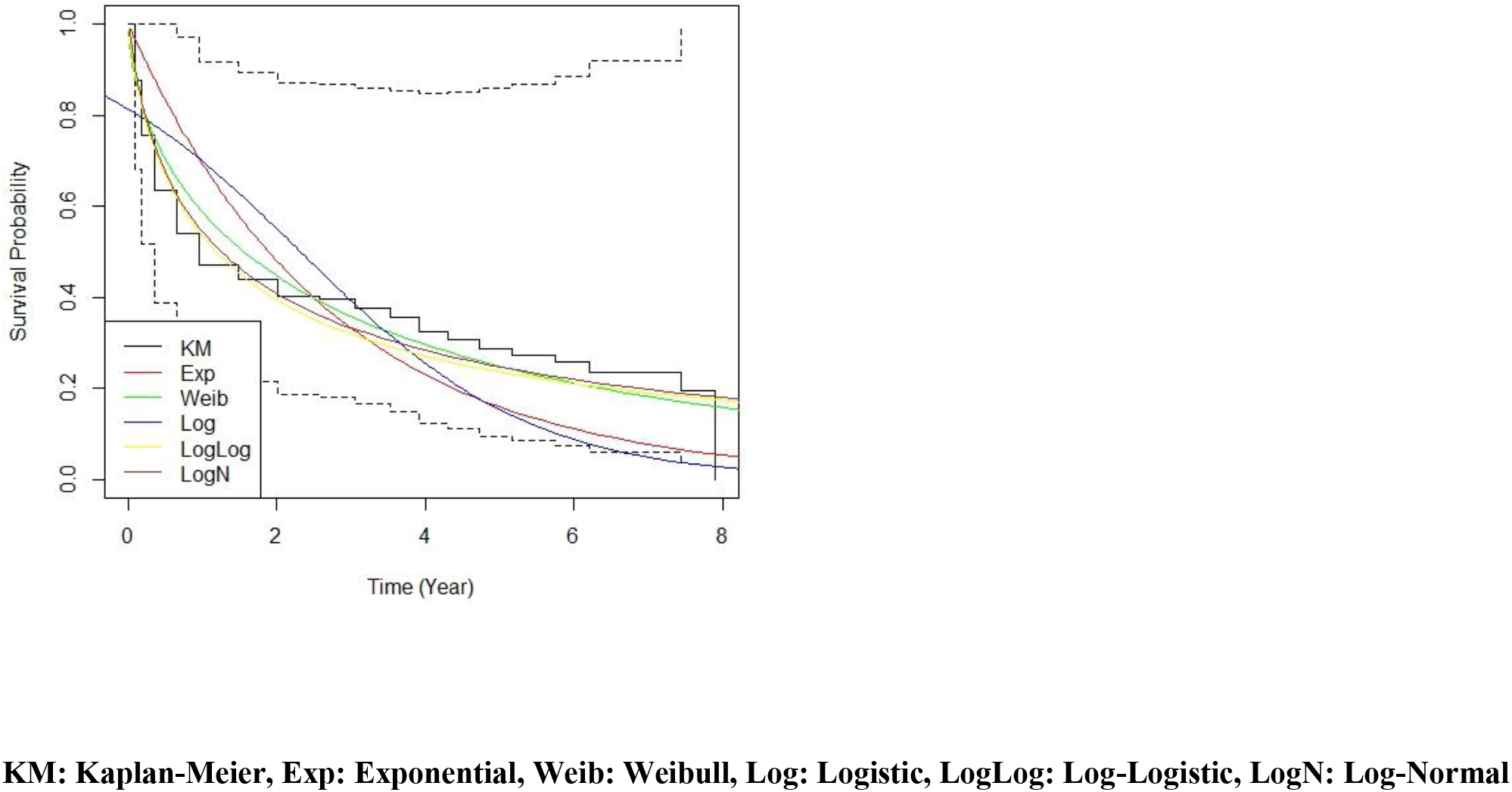
Estimated OS curves for the CPC 4 using different distributions.

**Table A1:** Estimated parametric Weibull survival functions are presented

|  |  |  |
| --- | --- | --- |
| Survival parameters for CPC1 | Weibull | Value |
|  | Mean | 25.70 |
|  | intercept | 3.24 |
|  | log(scale) | 0.26 |
|  | lambda | 0.08 |
|  | gamma | 0.76 |
| Survival parameters for CPC2 | Weibull | Value |
|  | Mean | 8.11 |
|  | intercept | 2.09 |
|  | log(scale) | 0.30 |
|  | lambda | 0.21 |
|  | gamma | 0.73 |
| Survival parameters for CPC3 | Weibull | Value |
|  | Mean | 2.74 |
|  | intercept | 1.01 |
|  | log(scale) | 0.22 |
|  | lambda | 0.44 |
|  | gamma | 0.80 |
| Survival parameters for CPC4 | Weibull | Value |
|  | Mean | 0.61 |
|  | intercept | -0.49 |
|  | log(scale) | 0.56 |
|  | lambda | 1.32 |
|  | gamma | 0.56 |

### Appendix II Result of cost-effectiveness analysis over hospital discharge horizon

The results of the base-case analysis with a short-term time horizon for Thermogard XP versus comparators showed that the total discounted cost per patient for the intervention was £26,469. In comparison, the total discounted cost for Blanketrol III and Arctic sun 500 was £26,039 and 26,625 respectively. Treatment with Thermogard XP led to an increase of 0.01 in discounted QALYs relative to Blanketrol III and Arctic sun 500 (*Table A2*). The estimated incremental cost-effectiveness ratio (ICER) per QALY gained was £48,531 versus Blanketrol III over hospital discharge duration and the ICER was dominant for Thermogard XP versus Arctic sun 500 over the hospital discharge duration. The results of the probabilistic sensitivity analysis are shown in *Figures A5* and *A5b* in the form of CEAC plots of Thermogard XP compared with Blanketrol III and Arctic sun 5000 respectively. If the hypothetical willingness-to-pay threshold for a QALY were £20,000, the probability of being cost-saving for TGXP would be 31 % and 54.6% versus Blanketrol II and Arctic sun 500 respectively.

**Table 2:** Result of cost-effectiveness analysis over hospital discharge horizon.

| Thermogard XP | Thermogard XP | Surface cooling | Incremental |
| --- | --- | --- | --- |
| Cost |  |  |  |
| Average costs per patient *versus Blanketrol III* | £26,469 | £26,039 | £430 |
| Average costs per patient *versus Arctic Sun 5000* | £26,469 | £26,625 | (£156) |
| Effectiveness |  |  |  |
| Total QALYs lived per patient | 0.11 | 0.10 | 0.01 |
| ICER *versus Blanketrol III* | £48,531 |  |  |
| ICER *versus Arctic Sun 5000* | Dominant |  |  |
| Probability of being cost-saving (at £20,000 WTP) *versus Blanketrol III* | 31% |  |  |
| Probability of being cost-saving (at £20,000 WTP) *versus Arctic Sun 5000* | 54.6% |  |  |

**Figure 5.**
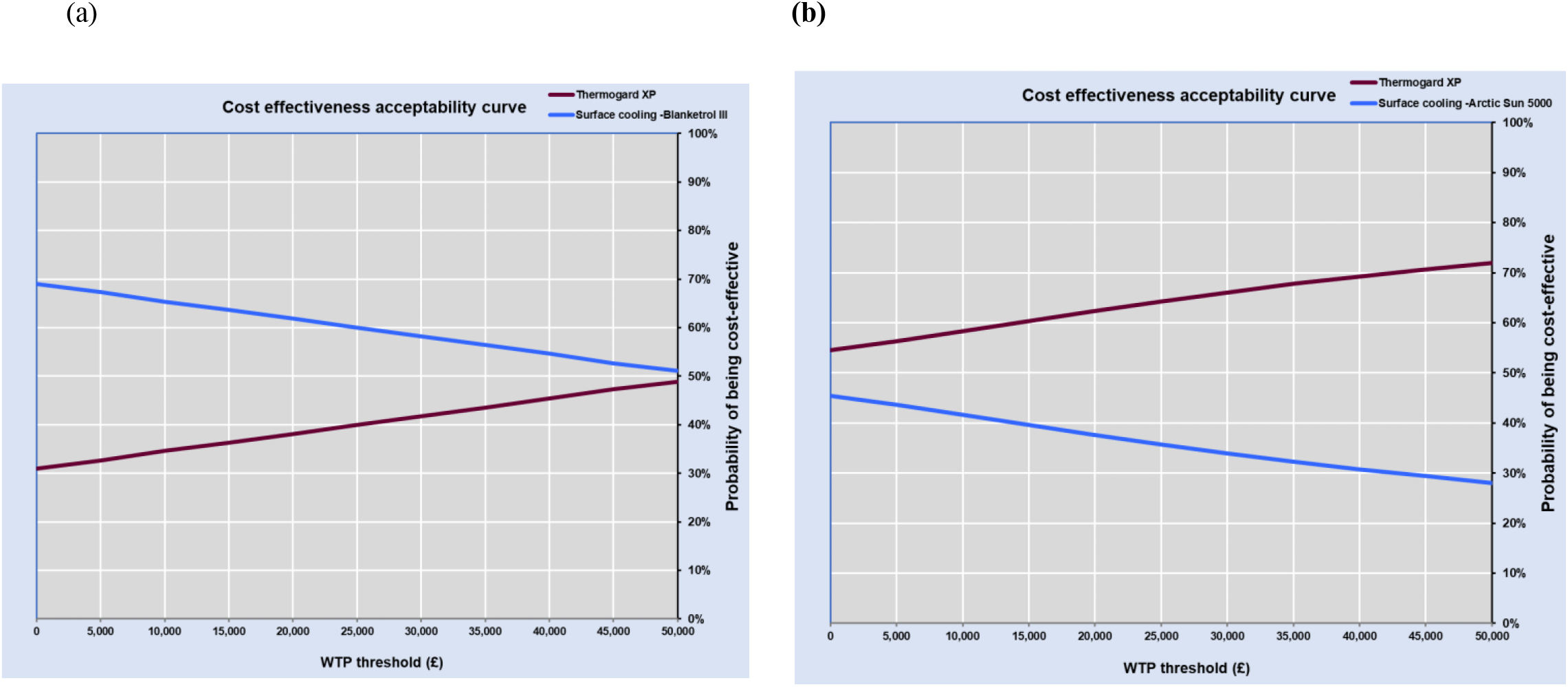
Cost-effectiveness acceptability curve plot of Thermogard XP compared with Blanketrol III (a) and Arctic sun 5000 (b)

## References

1. Overview | Therapeutic hypothermia following cardiac arrest | Guidance | NICE. Accessed August 7, 2020. https://www.nice.org.uk/guidance/ipg386

2. Vaity C, Al-Subaie N, Cecconi M. Cooling techniques for targeted temperature management post-cardiac arrest. Crit Care. 2015;19(1). doi:10.1186/s13054-015-0804-1

3. Laver S, Farrow C, Turner D, Nolan J. Mode of death after admission to an intensive care unit following cardiac arrest. Intensive Care Med. 2004;30(11):2126–2128. doi:10.1007/s00134-004-2425-z

4. Hawkes C, Booth S, Ji C, et al. Epidemiology and outcomes from out-of-hospital cardiac arrests in England. Resuscitation. 2017;110:133–140. doi:10.1016/j.resuscitation.2016.10.030

5. Thomassen A, Wernberg M. Prevalence and Prognostic Significance of Coma after Cardiac Arrest Outside Intensive Care and Coronary Units. Acta Anaesthesiol Scand. 1979;23(2):143–148. doi:10.1111/j.1399-6576.1979.tb01434.x

6. Bartlett ES, Valenzuela T, Idris A, et al. Systematic review and meta-analysis of intravascular temperature management vs. surface cooling in comatose patients resuscitated from cardiac arrest. Resuscitation. 2020;146:82–95. doi:10.1016/j.resuscitation.2019.10.035

7. (23) (PDF) Efficacy of different cooling technologies for therapeutic temperature management: A prospective intervention study. ResearchGate. Accessed August 11, 2020. https://www.researchgate.net/publication/322073031_Efficacy_of_different_cooling_technologies_for_therapeutic_temperature_management_A_prospective_intervention_study

8. Summary | Thermogard XP for therapeutic hypothermia after cardiac arrest | Advice | NICE. Accessed August 7, 2020. https://www.nice.org.uk/advice/mib37/chapter/summary

9. Methods for the Economic Evaluation of Health Care Programmes - Research Database, The University of York. Accessed August 7, 2020. https://pure.york.ac.uk/portal/en/publications/methods-for-the-economic-evaluation-of-health-care-programmes(8f69bcee-cdac-44fa-871c-f821470df60a).html

10. guide-to-the-methods-of-technology-appraisal-2013-pdf-2007975843781.pdf. Accessed August 7, 2020. https://www.nice.org.uk/process/pmg9/resources/guide-to-the-methods-of-technology-appraisal-2013-pdf-2007975843781

11. Nolan JP, Sandroni C, Böttiger BW, et al. European Resuscitation Council and European Society of Intensive Care Medicine Guidelines 2021: Post-resuscitation care. Resuscitation. 2021;161:220–269. doi:10.1016/j.resuscitation.2021.02.012

12. Calabró L, Bougouin W, Cariou A, et al. Effect of different methods of cooling for targeted temperature management on outcome after cardiac arrest: a systematic review and meta-analysis. Crit Care Lond Engl. 2019;23(1):285. doi:10.1186/s13054-019-2567-6

13. Phelps R, Dumas F, Maynard C, Silver J, Rea T. Cerebral Performance Category and long-term prognosis following out-of-hospital cardiac arrest. Crit Care Med. 2013;41(5):1252–1257. doi:10.1097/CCM.0b013e31827ca975

14. Improved curve fits to summary survival data: application to economic evaluation of health technologies | BMC Medical Research Methodology | Full Text. Accessed August 23, 2019. https://bmcmedresmethodol.biomedcentral.com/articles/10.1186/1471-2288-11-139

15. Merchant Raina M., Becker Lance B., Abella Benjamin S., Asch David A., Groeneveld Peter W. Cost-Effectiveness of Therapeutic Hypothermia After Cardiac Arrest. Circ Cardiovasc Qual Outcomes. 2009;2(5):421–428. doi:10.1161/CIRCOUTCOMES.108.839605

16. Petrie J, Easton S, Naik V, Lockie C, Brett SJ, Stümpfle R. Hospital costs of out-of-hospital cardiac arrest patients treated in intensive care; a single centre evaluation using the national tariff-based system. BMJ Open. 2015;5(4). doi:10.1136/bmjopen-2014-005797

17. The technology | Arctic Sun 5000 for therapeutic hypothermia after cardiac arrest | Advice | NICE. Accessed August 11, 2020. https://www.nice.org.uk/advice/mib112/chapter/The-technology

18. Chan Paul S., McNally Bryan, Nallamothu Brahmajee K., et al. Long-Term Outcomes Among Elderly Survivors of Out-of-Hospital Cardiac Arrest. J Am Heart Assoc. 5(3):e002924. doi:10.1161/JAHA.115.002924

19. NHS reference costs 2015 to 2016. GOV.UK. Accessed August 28, 2019. https://www.gov.uk/government/publications/nhs-reference-costs-2015-to-2016

20. Stiell Ian, Nichol Graham, Wells George, et al. Health-Related Quality of Life Is Better for Cardiac Arrest Survivors Who Received Citizen Cardiopulmonary Resuscitation. Circulation. 2003;108(16):1939–1944. doi:10.1161/01.CIR.000009502895929.B0

21. Hurdus B, Munyombwe T, Dondo TB, et al. Association of cardiac rehabilitation and health-related quality of life following acute myocardial infarction. Heart. 2020;106(22):1726–1731. doi:10.1136/heartjnl-2020-316920

22. Fryback DG, Dunham NC, Palta M, et al. U.S. NORMS FOR SIX GENERIC HEALTH-RELATED QUALITY-OF-LIFE INDEXES FROM THE NATIONAL HEALTH MEASUREMENT STUDY. Med Care. 2007;45(12):1162–1170. doi:10.1097/MLR.0b013e31814848f1

23. Gage BF, Cardinalli AB, Owens DK. The effect of stroke and stroke prophylaxis with aspirin or warfarin on quality of life. Arch Intern Med. 1996;156(16):1829–1836.

24. Raina Ketki D, Callaway Clifton, Rittenberger Jon C, Holm Margo B. Neurological and functional status following cardiac arrest: Method and tool utility. Resuscitation. 2008;79(2):249–256. doi:10.1016/j.resuscitation.2008.06.005

25. Gillies MA, Pratt R, Whiteley C, Borg J, Beale RJ, Tibby SM. Therapeutic hypothermia after cardiac arrest: A retrospective comparison of surface and endovascular cooling techniques. Resuscitation. 2010;81(9):1117–1122. doi:10.1016/j.resuscitation.2010.05.001

26. Tømte Ø, Drægni T, Mangschau A, Jacobsen D, Auestad B, Sunde K. A comparison of intravascular and surface cooling techniques in comatose cardiac arrest survivors. Crit Care Med. 2011;39(3):443–449. doi:10.1097/CCM.0b013e318206b80f

27. Deye N, Cariou A, Girardie P, et al. Endovascular Versus External Targeted Temperature Management for Patients With Out-of-Hospital Cardiac Arrest: A Randomized, Controlled Study. Circulation. 2015;132(3):182–193. doi:10.1161/CIRCULATIONAHA.114.012805

28. Glover GW, Thomas RM, Vamvakas G, et al. Intravascular versus surface cooling for targeted temperature management after out-of-hospital cardiac arrest – an analysis of the TTM trial data. Crit Care. 2016;20(1):381. doi:10.1186/s13054-016-1552-6

29. Home. NHS Supply Chain. Accessed July 8, 2021. https://www.supplychain.nhs.uk/

30. Look X, Li H, Ng M, et al. Randomized controlled trial of internal and external targeted temperature management methods in post-cardiac arrest patients. Am J Emerg Med. 2018;36(1):66–72. doi:10.1016/j.ajem.2017.07.017

31. Oh SH, Oh JS, Kim Y-M, et al. An observational study of surface versus endovascular cooling techniques in cardiac arrest patients: a propensity-matched analysis. Crit Care. 2015;19(1). doi:10.1186/s13054-015-0819-7

32. Pittl U, Schratter A, Desch S, et al. Invasive versus non-invasive cooling after in-and out-of-hospital cardiac arrest: a randomized trial. Clin Res Cardiol. 2013;102(8):607–614. doi:10.1007/s00392-013-0572-3

33. Sonder P, Janssens GN, Beishuizen A, et al. Efficacy of different cooling technologies for therapeutic temperature management: A prospective intervention study. Resuscitation. 2018;124:14–20. doi:10.1016/j.resuscitation.2017.12.026

34. Ferreira IA, Schutte M, Oosterloo E, et al. Therapeutic mild hypothermia improves outcome after out-of-hospital cardiac arrest. Neth Heart J. 2009;17(10):378–384. doi:10.1007/BF03086288

35. Kim KH, Shin SD, Song KJ, et al. Cooling methods of targeted temperature management and neurological recovery after out-of-hospital cardiac arrest: A nationwide multicenter multi-level analysis. Resuscitation. 2018;125:56–65. doi:10.1016/j.resuscitation.2018.01.043

36. Waard MC de, Banwarie RP, Jewbali LSD, Struijs A, Girbes ARJ, Groeneveld ABJ.Intravascular versus surface cooling speed and stability after cardiopulmonary resuscitation. Emerg Med J. 2015;32(10):775–780. doi:10.1136/emermed-2014-203811

37. Flint AC, Hemphill JC, Bonovich DC. Therapeutic Hypothermia after Cardiac Arrest: Performance Characteristics and Safety of Surface Cooling with or without Endovascular Cooling. Neurocrit Care. 2007;7(2):109–118. doi:10.1007/s12028-007-0068-y

38. Flemming: Comparison of external and intravascular… - Google Scholar. Accessed August 8, 2020. https://scholar.google.com/scholar_lookup?title=Comparison%20of%20external%20and%20intravascular%20cooling%20to%20induce%20hypothermia%20in%20patients%20after%20CPR&journal=Ger%20Med%20Sci&volume=4&publication_year=2006&author=Flemming%2CK&author=Simonis%2CG&author=Ziegs%2CE

39. Forkmann M, Kolschmann S, Holzhauser L, et al. Target temperature management of 33°C exerts benefi cial haemodynamic eff ects after out-of-hospital cardiac arrest. Acta Cardiol. 2015;70(4):451–459. doi:10.1080/AC.70.4.3096893

40. Rosman J, Hentzien M, Dramé M, et al. A comparison between intravascular and traditional cooling for inducing and maintaining temperature control in patients following cardiac arrest. Anaesth Crit Care Pain Med. 2018;37(2):129–134. doi:10.1016/j.accpm.2016.08.009

41. Gajarski RJ, Smitko K, Despres R, Meden J, Hutton DW. Cost-effectiveness analysis of alternative cooling strategies following cardiac arrest. SpringerPlus. 2015;4:427. doi:10.1186/s40064-015-1199-9

42. Kirk A, McDaniel C, Szarlej D, Rincon F. Assessment of Antishivering Medication Requirements During Therapeutic Normothermia: Effect of Cooling Methods. Ther Hypothermia Temp Manag. 2016;6(3):135–139. doi:10.1089/ther.2016.0001

43. Taccone FS, Picetti E, Vincent J-L. High Quality Targeted Temperature Management (TTM) After Cardiac Arrest. Crit Care Lond Engl. 2020;24(1):6. doi:10.1186/s13054-019-2721-1

44. De Fazio C, Skrifvars MB, Søreide E, et al. Intravascular versus surface cooling for targeted temperature management after out-of-hospital cardiac arrest: an analysis of the TTH48 trial. Crit Care. 2019;23(1):61. doi:10.1186/s13054-019-2335-7

45. National life tables: UK - Office for National Statistics. Accessed March 15, 2021. https://www.ons.gov.uk/peoplepopulationandcommunity/birthsdeathsandmarriages/lifeexpectancies/datasets/nationallifetablesunitedkingdomreferencetables

